# Estimating the likelihood of epilepsy from clinically non-contributory EEG using computational analysis: A retrospective, multi-site case-control study

**DOI:** 10.1101/2023.03.08.23286937

**Authors:** Luke Tait, Lydia E Staniaszek, Elizabeth Galizia, David Martin-Lopez, Matthew C Walker, Al Anzari Abdul Azeez, Kay Meiklejohn, David Allen, Chris Price, Sophie Georgiou, Manny Bagary, Sakh Khalsa, Francesco Manfredonia, Phil Tittensor, Charlotte Lawthom, Rohit Shankar, John R Terry, Wessel Woldman

## Abstract

**Background:** A retrospective, multi-site case control study was carried out to validate a set of candidate biomarkers of seizure susceptibility. The objective was to determine the robustness of these biomarkers derived from routinely collected EEG within a large cohort (both epilepsy and common alternative conditions which may present with a possible seizure, such as NEAD).

**Methods:** The database consisted of 814 EEG recordings from 648 subjects, collected from 8 NHS sites across the UK. Clinically non-contributory EEG recordings were identified by an experienced clinical scientist (N = 281; 152 alternative conditions, 129 epilepsy). Eight computational markers (spectral [N = 2], network-based [N = 4] and model-based [N = 2]) were calculated within each recording. Ensemble-based classifiers were developed using a two-tier cross-validation approach. We used standard regression methods in order to identify whether potential confounding variables (e.g. age, gender, treatment-status, comorbidity) impacted model performance.

**Findings:** We found levels of balanced accuracy of 68% across the cohort with clinically non-contributory normal EEGs (sensitivity: 61%, specificity: 75%, positive predictive value: 55%, negative predictive value: 79%, diagnostic odds ratio: 4.64). Group-level analysis found no evidence suggesting any of the potential confounding variables significantly impacted the overall performance.

**Interpretation:** These results provide evidence that the set of biomarkers could provide additional value to clinical decision-making, providing the foundation for a decision support tool that could reduce diagnostic delay and misdiagnosis rates. Future work should therefore assess the change in diagnostic yield and time to diagnosis when utilising these biomarkers in carefully designed prospective studies.

**Research in Context:** *Evidence before this study:* We searched Google Scholar and Pubmed (March 21, 2022) for the following phrases ((“EEG” OR “electroencephalogram” OR “electroencephalography”) AND (“biomarker”) AND (“epilepsy” OR “seizure”) AND (“resting state” OR “resting-state”) OR (“normal”)). Several of the existing studies developed deep learning approaches for identifying the presence of interictal epileptiform discharges (IED), with the overarching aim to develop an automated stand-alone diagnostic tool. These approaches are particularly sensitive to the potential presence of artefacts in the EEG recordings and typically include spectral rather than network- or model-based features. We found no studies of more than 100 participants that assessed the cross-validated performance of candidate biomarkers on routine EEG recordings that were clinically non-contributory. One study found near-chance performance of a deep-learning based method using spectral features on a smaller cohort of people suspected of epilepsy (N=33 epilepsy; N=30 alternative conditions) with clinically non-contributory EEGs. Another study found overall accuracy of 69% (N=74 epilepsy; N=74 alternative conditions) but this framework did not use any independent cross-validation methods. Estimates of sensitivity of clinical markers of seizure susceptibility in routine EEG recordings vary between 17-56%. To the best of our knowledge no studies have assessed whether computational biomarkers offer sufficient discrimination between people with epilepsy and an alternative diagnosis to provide potential decision support for people with suspected epilepsy.

*Added value of this study:* We show that data-driven analysis of routinely collected EEGs that are currently considered clinically non-informative (i.e. absence of apparent epileptiform activity) can be used to distinguish EEGs from people with epilepsy from people with an alternative diagnosis with better-than-chance performance. To the best of our knowledge, this is the largest retrospective study assessing the performance of computational biomarkers derived from clinically non-contributory EEG recordings. The resulting statistical model is interpretable and relies on both spectral and computational (network- and model-based) features. We perform a series of validity and sensitivity analysis to assess the overall robustness of the final statistical model used for classification. We also conduct several statistical tests to analyse any shared characteristics (e.g. site, comorbidity) amongst the primary classes (FP, FN, TP, TN). These findings validate previous biomarker discovery- or development-studies, and provide evidence that they offer better-than-chance performance in a clinically relevant context. Future large-scale studies could consider combining these methods with interictal features for non-specialist settings.

*Implications of all the available evidence:* Our study presents evidence that computational analysis of clinically non-contributory EEGs could provide additional decision support for both epilepsy and alternative conditions. Since the statistical model and underlying features are interpretable, they could provide the starting point for further exploring the mechanisms that drive overall seizure-likelihood. Future work should focus on prospective testing and validation (e.g. identification of specific situations or cases in which these methods could be of added value) as well as assessing heterogeneity across different syndromes and diagnoses (e.g. NEAD, focal vs generalised epilepsy).

## Introduction

Epilepsy affects over 50 million people worldwide, with estimates suggesting around 2.4 million new cases worldwide per year.^1^ Epilepsy remains a clinical diagnosis, based on expert analysis of likelihood of further seizures, a decision which considers multiple factors including a person’s medical history, and results from routine diagnostic tests such as the scalp electroencephalogram (EEG). However, the sensitivity of routine EEG in the identification of persons with epilepsy remains low relying on expert identification of interictal epileptiform discharges (IED).^2,3^ The overall specificity of routine EEG is broadly estimated to fall within a range of 78-98%, but individual studies often report wide confidence intervals.^2,3^ Additionally, it is currently recommended that EEGs which lack IED (herein termed “non-contributory EEGs”) should not be used in isolation to exclude a diagnosis of epilepsy (see e.g. NICE guidelines 2017).^4^ As a result, delay in both the diagnosis of epilepsy and its differentials is common, driving research into identification of biomarkers of epilepsy, using routine EEG as a substrate. Computational approaches to interrogate routine EEG have attracted much interest recently. Typically, these have focussed on automatic identification of IED, and / or computation of whole-brain networks by analysis of resting state EEG, that is, those portions of EEG in which no epileptiform features are present.^5-7^ Automatic identification of IED is both sensitive to EEG artefacts and its performance is limited by the levels of identification achieved by the trained expert.

In contrast, approaches that do not require the presence of IED offer the potential to improve sensitivity of routine EEG, since they focus on features that are not currently considered clinically informative. For example, computational analysis of resting-state EEG has consistently revealed differences in whole-brain network measures at the group level. These differences, confirmed by meta-analysis, have been shown in case-controlled studies in both generalised and focal epilepsies when compared to healthy participants.^6,8^ Whilst constituting Phase I – level evidence, it is unclear whether these findings are translatable to patient cohorts typical to that seen in clinical practice. Few studies have assessed group-level changes in resting-state or clinically non-contributory EEG between persons with epilepsy and persons with an alternative condition (e.g. syncope or non-epileptic attack disorder (NEAD)). However, those that did have also confirmed the presence of identifiable group level differences, demonstrating the potential of computational biomarkers both in the presence of identifiable IED, and in non-contributory EEGs.^9^ It is also unclear how group level effects translate to the individual level. For example, the group level differences observed in Larsson and colleagues were shown to offer very limited predictive capacity at the individual level.^5,10,11^ A further limitation in the studies described above is that typically they were performed on data obtained from a small number of diagnostic centres.

Addressing these challenges, we aimed to develop a robust classification pipeline to validate a set of candidate biomarkers in a way that is relevant to clinical practice. To this end, we performed a retrospective, multi-site case control study. Our study includes people ultimately diagnosed with epilepsy, as well as those ultimate diagnosed with common alternatives. To maximise robustness and increase its clinical applicability, we leveraged a two-tier cross-validation approach, and made no exclusions based on comorbidity (including neurological), medication (including anti-seizure medications or medications with known effects on the EEG), or time since first or most recent seizure or seizure-like event.

## Methods

### Study Design and Participants

Eight sites within the NHS participated in this study. Inclusion and exclusion criteria may be found at clinicaltrials.gov (Identifier: NCT05384782). In summary, inclusion requirements were as follows: The subject was suspected of having had a seizure or epilepsy, and as part of the diagnostic process one or more EEGs were recorded with a minimum of 19 channels, applied to the 10-20 international system of electrode placement. The subject ended up with a confirmed diagnosis of epilepsy or of a common alternative condition such as syncope, or non-epileptic attack disorder with the confirmed clinical diagnosis remaining stable for at least one year. Additional meta-data including patient age, sex, comorbidities, anti-seizure medication status, EEG result were also provided (appendix p1).

EEGs which were reported by the consultant neurophysiologist of each participating site as non-contributory were included in this study. Both EEGs that were classed as “normal” and “abnormal” were included. These labels were compared with the EEG for consistency by an experienced clinical scientist (LS). For classification purposes an “abnormal” EEG included EEGs which contained abnormal features which were not specific for epilepsy, and which therefore did not contribute to the ultimately confirmed diagnosis. An example of such an EEG could include incidental findings of non-specific abnormalities such as those which may be of a vascular, pharmacological, structural or metabolic pathophysiological origin. Due to the inherent heterogeneity introduced by the presence or absence of EEG abnormalities, individual classifiers will be developed for the normal and the abnormal clinically non-contributory cohorts separately.

### EEG Pre-processing

Scalp EEG data were imported into Matlab (R2021b). The same nineteen clinical EEG channels (see appendix p1) were chosen for all participants across all sites, while any other channels were discarded. Recordings were notch filtered at 50 Hz and bandpass filtered from 0.53-70 Hz using a 4^th^ order Butterworth zero-phase filter and re-referenced to average.

### Candidate Markers

Published resting-state biomarkers of EEG were reviewed in the literature making use of Google Scholar, PubMed. We searched for papers which had reported areas under ROC curves (AUROC) for statistical models classifying epilepsy from resting-state EEG/MEG and used these AUROC values as the a-priori effect size. A total of six papers were identified, with AUROC values ranging from 0.43-0.99.^12-17^ Following the method of Riley and colleagues, we found seven estimates for the maximal number of markers.^18^ Of these seven estimates, six suggested a maximum of eight markers and one suggested a maximum of seven markers. Hence, we proceeded to select the first eight markers from an ordered list of candidate markers (by individual effect-size).^5^

#### Frequency-based markers

Two frequency-based markers were calculated. The peak-alpha frequency (PAF) was calculated by averaging the power spectrum of the occipital electrodes (O1 and O2) and calculating the frequency corresponding to the peak of the spectrum in the 8-13 Hz range.^17,19,20^ The alpha power (AP) was determined by calculating the 10.5-13.5 Hz relative power (averaged across channels after bipolar-referencing of the frontal and temporal channels.^15^

#### Network-based markers

The low-alpha band (6-9 Hz) phase-locking value (PLV) was computed between pairs of electrodes, and four graph theoretical markers were calculated from these weighted undirected networks.^21,22^ These markers were mean degree (MD), degree variance (DV), average weighted clustering coefficient (AWCC) and characteristic path length (CPL).^23^

#### Model-based markers

The final two markers are derived from the phenomenological multi-scaled oscillator model of the EEG.^11,12^ For both markers, an individual’s low-alpha PLV network and alpha power are used to parameterise a model which simulates EEG in which each electrode is described as a system of coupled oscillators (locally coupled according to each channel’s alpha power and scaled with a value called ‘local coupling’), and electrodes are connected according to the PLV weights (scaled by some value called ‘global coupling’). The first marker, critical coupling (CC), is the theoretical value of global coupling which causes the oscillators to synchronise (i.e. a simulated seizure). The second marker, local coupling (LC), is the theoretical value of local coupling (for a single simulated EEG electrode) which causes synchronisation. The marker corresponds to the maximum value across the 19 channels (representing the single most ‘ictogenic’ node in the model).

#### Marker calculation

All markers were implemented in Matlab (R2021b) using built-in Matlab functions and toolboxes (e.g. the signal-processing toolbox for frequency-based markers, https://www.mathworks.com/products/signal.html), the brain connectivity toolbox and previously published scripts for model-based markers.^12,23^ To minimise the effect of artefacts or non-specific abnormalities, EEG data was segmented into 20s epochs and the eight markers were calculated for each epoch. The median value for each marker across all 20s epochs was subsequently used for further analysis. Within each recording, 20 minutes of EEG was selected. The chosen starting time being sampled from the distribution of starting times of all routine EEGs within the study.

### Confounder model

Markers were adjusted for effects of confounders such as age, comorbidities, anti-seizure medications (ASM), and sex. To do so, we constructed a generalised linear model of the form:

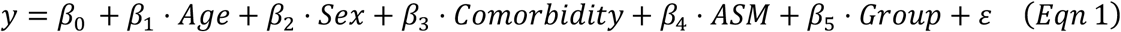

where *y* is a given marker; *Age* is participant age; *Sex, Comorbidity, ASM*, and

Group each have a value equal to one if the participant is respectively male, has comorbid disorders (e.g. dementia, stroke, etc), is being treated with anti-seizure medications, and has epilepsy or have value zero otherwise; *β*_*i*_, *i* ∈ {0, …, 5} are the parameters of the model; and *ε* are the participant-specific residuals. Prior to linear modelling, the relevant variables were transformed using a monotonic function (e.g. log transform) to better approximate normality in the residuals, a key assumption of linear models. Note that during hold-out cross-validation confounder correction was applied to both training and hold-out sets using linear models fitted to the training set (i.e. the model in Equation 1 used only training data to estimate the *β*^*i*^ coefficients, independently of hold-out set).

### Statistical Model

Data was partitioned using 10-fold cross validation, and a suite of statistical models were trained using Matlab’s ‘*Classification Learner’* application (figure 2). For each fold, prior to training principal component analysis (PCA) was performed on the training set to reduce the eight confounder-corrected markers, and the PCA weights from the training set were subsequently applied to the hold-out set. A full description of the hyperparameters for model training is given in the appendix (p1). Model performance was characterised using a set of well-established outcome metrics including balanced accuracy, AUROC, and Brier score (see appendix p3). Potential relationships between confounders and overall performance were tested with Fisher’s exact test (95% significance) and the Kruskal-Wallis test (95% significance), using Bonferroni correction for multiple comparisons.

**Figure 1:**
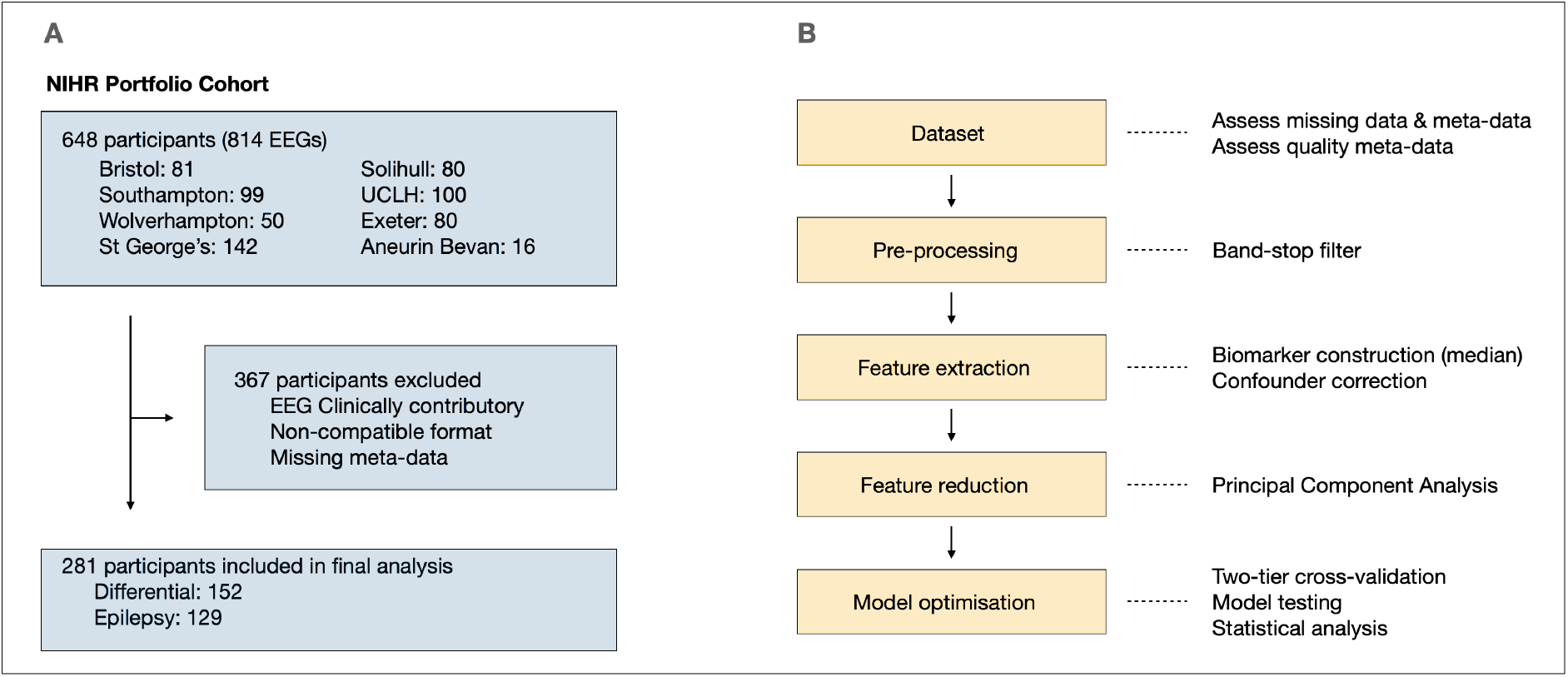
Cohort Profile & Analysis Pipeline. **Caption**: A: Cohort profile: final analysis was carried out on the first available EEG of each participant that was clinically non-contributory. B: flow diagram summarising analysis steps.

**Figure 2:**
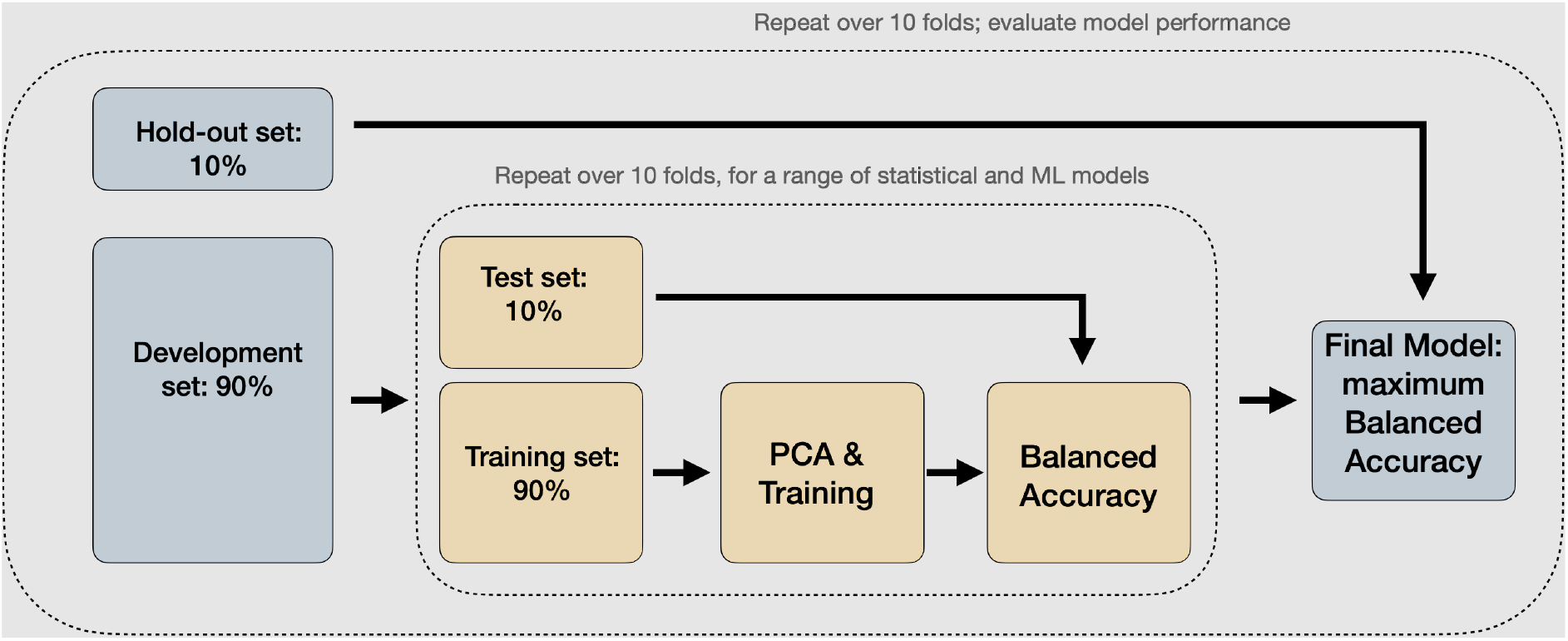
Two-tier cross-validation. **Caption**: Flow diagram depicting the two-tier cross-validation scheme for model development. Each individual EEG is part of a hold-out set exactly once. A suite of statistical models is explored to find a model with optimal performance in terms of balanced accuracy during the training phase. Then this model is tested against the independent hold-out sets. Balanced accuracy is reported for the cross-validated development set and the final hold-out test-set. Full details on the modelling steps are given in the appendix (p1-2).

## Results

Retrospective screening took place between December 1, 2019 and March 31, 2022 at a total of 8 clinical sites. If a participant was included for the study, all available EEG recordings were uploaded for each participant to a central data repository, along with their meta-data. A total of 814 EEG files from 648 individual patients were reported as uploaded. After participant recruitment, all meta-data were analysed by an experienced clinical scientist (LS). EEG results were classified as indicative of epilepsy, non-specifically abnormal (not including those indicative of epilepsy), or normal. Those classified as non-specifically abnormal or normal were defined as being clinically non-contributory. For each included participant, the first recorded non-contributory EEG was used. Participants were excluded from the present analysis if they had no non-contributory EEGs or if the EEG was provided in an unreadable data-format (see also ClinicalTrials.gov Identifier: NCT05384782).

The first recorded non-contributory EEG from 281 participants were analysed for model development. Of these, 89 (31.7%) were reported by the consultant neurophysiologist as non-specifically abnormal, and 192 (68.2%) as normal. Of the 281 subjects, 129 (45.9%) were ultimately diagnosed with epilepsy (9 generalised, 106 focal, and 14 unclassified – either unknown or information not available), and 152 (54.1%) were ultimately diagnosed with an alternative condition. Full details on age, gender, co-morbidity and treatment status are summarised in Table 1.

For the 192 normal non-contributory EEGs, the random undersampling boosting (RUB) model achieved the highest level of mean balanced accuracy (65.1%; s.d.: 3.0%) during ten-fold cross-validation in the training phase.^24^ The PCA approach reduced the feature space to four principal components (mean explained variance: 96.8%; s.d.: 0.12%). The final RUB model achieved balanced accuracy of 67.9% on the independent test-set (figure 3), with a diagnostic odds ratio of 4.6352 (95% C.I.: [2.4676, 8.7070]). An extensive set of performance metrics, including AUROC, recall, F1 score, Brier score and a confusion matrix, was calculated (see appendix p3).

**Figure 3:**
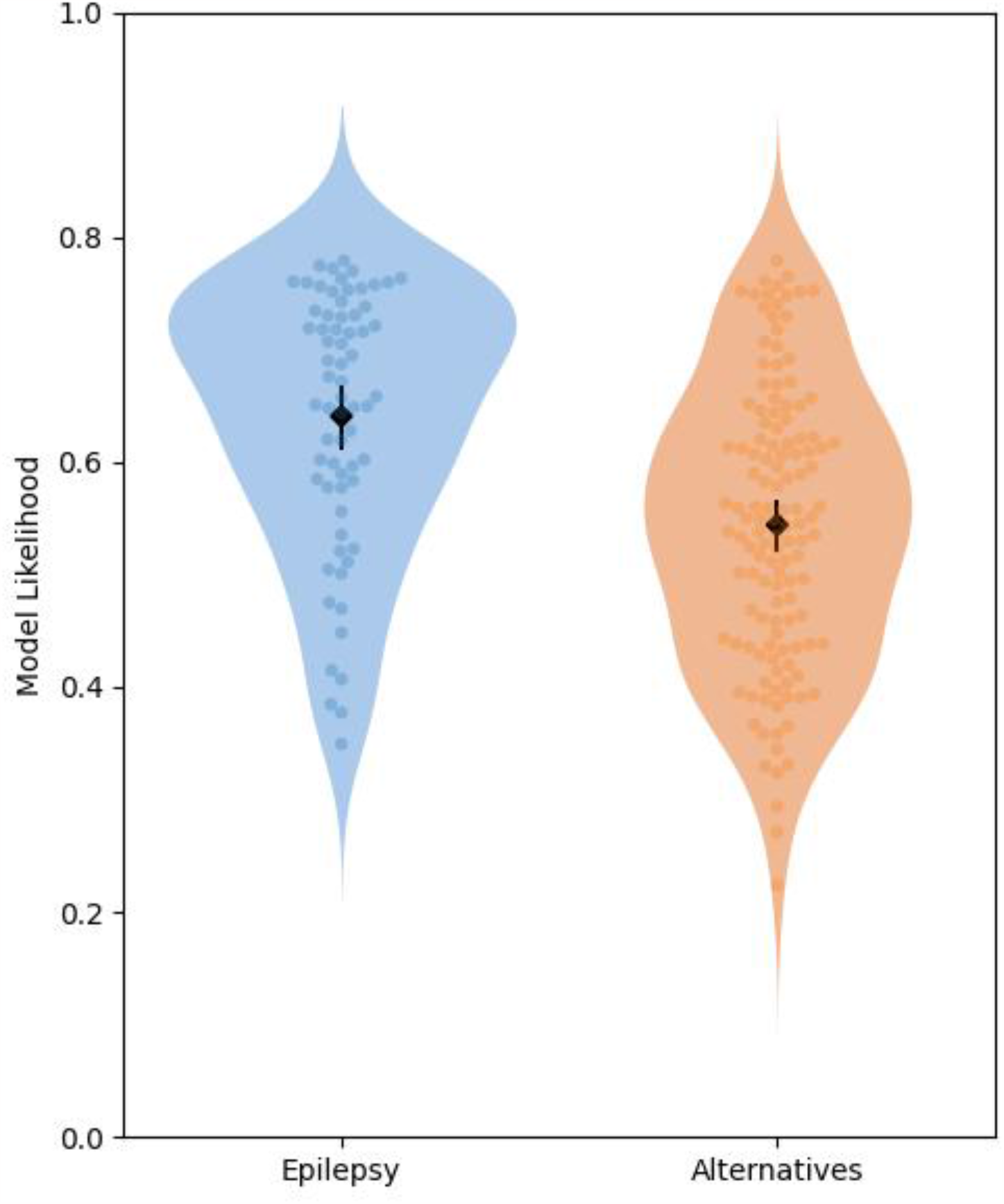
RUB likelihood scores for normal clinically non-contributory EEGs. **Caption:** violin plots displaying the likelihood scores from the random undersampling boosting (RUB) model across the epilepsy (blue) and alternative condition (orange) groups for whom their EEGs were considered clinically normal but non-contributory. Individual values for each EEG (subject) are displayed with a swarm plot (darker blue and orange). Mean (diamond) and 95% confidence intervals are displayed within each violin plot (black).

For the 89 abnormal non-contributory EEGs, the subspace discriminant (SD) model achieved the highest level of mean balanced accuracy (58.2%, s.d: 4.6%) during ten-fold cross-validation in the training phase. The PCA approach did not reduce the feature space to lower dimensionality. The ultimate SD model achieved balanced accuracy of 61.2% on the independent test-set (figure 4), and a diagnostic odds ratio of 2.4774 (95% C.I: [0.9430, 6.5081], see appendix p4 for an extensive list of performance metrics).

**Figure 4:**
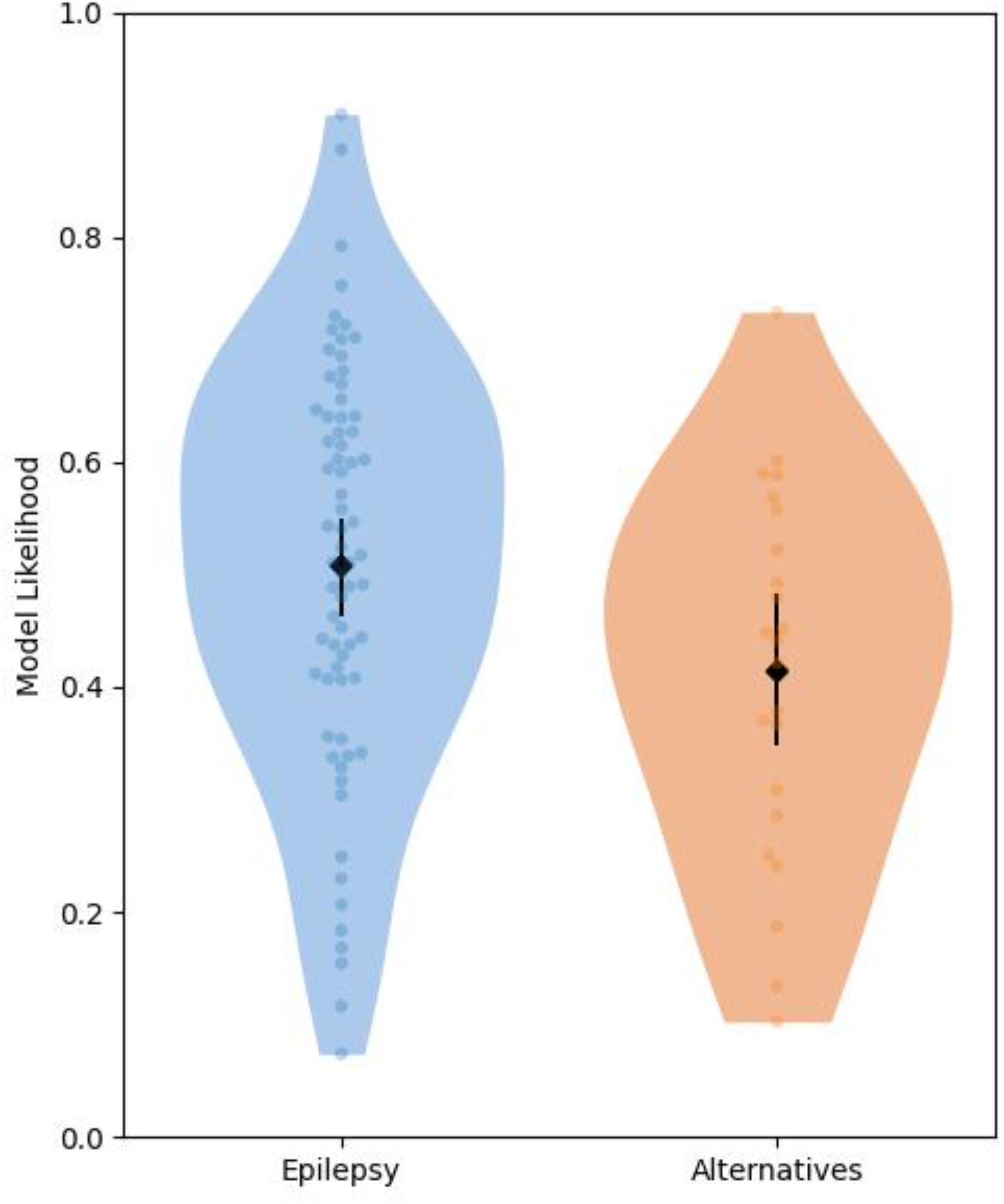
SD likelihood scores for abnormal clinically non-contributory EEGs. **Caption:** violin plots displaying the likelihood scores from the subspace discriminant (SD) model across the epilepsy (blue) and alternative condition(orange) groups for whom their EEGs were considered clinically abnormal but non-contributory. Individual values for each EEG (subject) are displayed with a swarm plot (darker blue and orange). Mean (diamond) and 95% confidence intervals are displayed within each violin plot (black).

At the level of the individual markers, several of the overall observed trends were concordant with the findings from the original development studies. In particular, there was an observed trend for increased mean degree, degree variance, average clustering coefficient and decreased characteristic path-length and critical coupling (appendix p5).

The discordant cases were further investigated. We found no association between misclassification and age, gender, ASM status, the presence of comorbidity, or specific clinical site (appendix p8).

## Discussion

In this retrospective, multi-site case-control study, clinically non-contributory EEG recordings were interrogated using a set of candidate biomarkers and a statistical model developed for classification. The models were cross-validated and tested on non-contributory EEGs from a cohort of 129 subjects with epilepsy and 152 subjects originally suspected of having epilepsy but ultimately receiving a diagnosis for an alternative condition. The models achieved increased performance in comparison to existing similar studies, contrasting favourably to the clinical yield for routine EEG recordings of this nature.^2,7^ Additionally, since the classifiers were tested on the first available non-contributory EEG recording for each subject without requiring clinical marking, the developed statistical model could potentially reduce diagnostic delay without increasing clinical workload.

The cohort of people enrolled in the study, both with epilepsy and with an alternative diagnosis, represent the variability of subjects expected in standard clinical practice in a way that single-site studies using healthy participants for control data cannot. Furthermore, in current clinical practice, given a contributory EEG (e.g. one with the presence of IED) can support a diagnosis of epilepsy, additional information gleaned from a digital biomarker is unlikely to alter clinical decision making or clinical management. The clinical biomarker will take precedent. Therefore, the true value in digital biomarkers of whole-brain networks lies in their ability to discriminate EEGs from persons with and without epilepsy when applied to non-contributory EEGs. Hence, to determine the potential added value of this set of candidate biomarkers, we assessed their performance only in first EEGs that were non-contributory. The routine clinical analysis of these recordings did not contribute one way or the other to a diagnosis at the time they were collected. This is a key differentiator of this study from other studies into candidate biomarkers for epilepsy, which typically include EEGs both containing IED and those without, or used high-density EEG.^5,7,25,26^ The approach presented here allowing the true added value of using biomarkers in addition to current standard clinical practice to be assessed. The results presented suggest that an existing set of candidate biomarkers that was evidenced on primarily Phase-I evidence, contained generalisable predictive power in a retrospective Phase-II setting.

The presence of non-specific abnormalities led to decreased performance for some key metrics, whereas other potential confounding variables (age, sex, co-morbidity, medication status, clinical site) did not specifically contribute to discordant cases. Interestingly, the overall decrease was primarily caused by decreased specificity, with very similar sensitivities, although the modest sample-size and class imbalance should be considered.

Our results may indicate that the non-specific abnormalities mediate the average properties of these biomarkers over the entirety of the EEG recording rather than having a pronounced, isolated effect at small sections of the EEGs. This finding suggest that the abnormal model might benefit from including additional non-linear, event-driven biomarkers as well as further studies that aim to specifically disambiguate the effect of local abnormalities on background properties.

Given the nature of the considered clinical cohort, this study has several limitations. First, all EEG recordings were collected from subjects of age 18 and above and therefore may not generalise to paediatric settings. Second, correcting for potential confounders with a basic regression model does not necessarily eliminate their full impact on either EEG features or comparisons. Third, even though this is to the best of our knowledge the largest study of its size (i.e. cross-validated, multi-site, clinically non-contributory EEG recordings), both statistical models were developed in unbalanced data-sets with relatively modest sample-sizes, in particular for the smallest class. Fourth, the separation between subjects as epilepsy or alternative does not map on directly to clinical decision making where the working diagnosis will ultimately be a specific condition or diagnosis (i.e. broad screening rather than a specific final diagnosis). Finally, there are a number of sources of potential variation that could not be controlled for. These include influence of recording time, subject alertness levels, or the choice of activation methods (e.g. intermittent photic stimulation, hyperventilation). These factors may have impacted key features within the EEG.

There are several important directions relevant to the future translation of these classification models. First, the models should be tested and verified against a large, independent retrospective data-set from at least one new clinical site. Next, the applicability of these markers to multiple classes should be tested and validated. Instead of separating subjects by ultimate diagnosis (epilepsy or alternative condition), these models should test and refine the biomarkers for a specific diagnosis or syndrome, such as NEAD or Juvenile Myoclonic Epilepsy (JME).^27^ The methodology could also explicitly consider the preference of clinicians in their intended use and context: for example, by prioritising specificity over sensitivity during the development of the algorithms. Another study should specifically consider the specific variability and variance found across clinical sites and within cohorts, including EEG durations (including longitudinal), activation methods, comorbidity and presence of (epileptiform) abnormalities. Finally, it would be particularly important to assess their impact on treatment decisions, for example at tertiary centres where people with suspected refractory epilepsy are investigated for potential surgery.

In conclusion, we have developed statistical models that may offer improved diagnostic yield for epilepsy, over and above the current clinical standard. This was achieved using a set of digital biomarkers derived from what would currently be considered clinically non-contributory EEG recordings. A promising direction for clinical translation could be the early identification of people with epilepsy after initial non-contributory EEGs for prolonged EEG monitoring, thereby reducing periods of watchful waiting and diagnostic delay. Future prospective and longitudinal studies are crucial to assess the overall utility of these digital markers for the purpose of diagnostic decision support.

## Data Availability

The EEG recordings and meta-data are not publicly available due to restrictions by privacy laws. Post-processed data supporting the findings of this study are available upon reasonable request from the corresponding author.

## Acknowledgements

LT was supported by the NIHR (AI01646).

WW was supported by Epilepsy Research UK (F2002), Innovate UK (103939) and the NIHR (AI01646).

JT was supported by EPSRC (EP/N014391/2 & EP/T027703/1), Innovate UK (103939), and the NIHR (AI01646).

RS was supported by Innovate UK (103939) and the NIHR (AI01646).

We thank Professor Sandor Beniczky for his comments on an earlier draft of the manuscript.

