## Supplementary Materials for "Estimating the likelihood of epilepsy from clinically non-contributory EEG using computational analysis: A retrospective, multi-site case-control study"

**Appendix**

**Meta-data**

| | Age | Sex | Treated with ASM | Presence of comorbidity |
|---|---|---|---|---|
| **Abnormal EEG** | Mean: 50.5 yrs; s.d.: 20.1 yrs; range: 18-91 yrs | 58 F (65.2%); 31 M (34.8%) | 40 (44.9%) | 64 (71.2%) |
| **Normal EEG** | Mean: 38.2 yrs; s.d.: 14.4 yrs; range: 18-91 yrs | 102 F (53.1%); 89 M (46.4%); 1 other (0.5%) | 70 (36.5%) | 123 (64.1%) |
| **Total** | Mean: 42.0 yrs; s.d.: 17.4 yrs; range: 18-91 yrs | 160 F (56.9%); 120 M (42.7); 1 other (0.4%) | 110 (39.2%) on ASM | 187 (64.8%) |

**EEG channels**

Fp1, Fp2, F3, F4, F7, F8, Fz, T3, T4, Cz, C3, C4, Pz, P3, P4, P7, P8, O1, O2.

**Choosing hyperparameters for model training**

The classification models presented in the main manuscript were ensemble-based (using '*fitcensemble*') and principal component analysis (PCA) was used to reduce the full set of markers to fewer dimensions. Here we describe the methodology used to select these hyperparameters to avoid overfitting to the data, as simply choosing the combination with optimal cross-validated accuracy is prone to overfitting.[1]

Data was partitioned using 10-fold cross validation. For each fold, 90% of the data was used as a development set, while 10% of the data was used as an independent hold-out set. Every participant was in the hold-out set for exactly one fold. Subsequently we analysed the development sets to choose the combination of classifier type and number of features. For all combinations of number of features (N=8) and the model types included in Matlab's Classification Learner App (https://mathworks.com/help/stats/classificationlearner-app.html) (only using default models and excluding optimizable models – a full list is given in the following section), we calculated 10-fold cross-validated balanced accuracy *within* the development set, i.e. we additionally partitioned the development set into 10 partitions, training the model on 90% of the development set and testing on 10% of the development set and repeating 10 times using each participant in the development set in exactly one test set and 9 training sets.

After choosing the model/number of dimensions with optimal cross-validated balanced accuracy within the development set, the classifier was retrained on the full development set. This model was subsequently used to predict responses for the hold-out set (the hold-out set was reduced to a lower dimensionality using the un-mixing weights from the PCA of the development set), which were then used to calculate the reported performance metrics. Hence, the hold-out set was not used in the choice of optimal model/dimensionality, minimising the risk of overfitting.

**List of models**

Listed below are the full set of classification models that were tested during the training phase. In all cases except logistic regression and neural networks (which do not support custom misclassification costs in Matlab's Classification Learner App), the misclassification costs were set inversely proportional to the number of participants in each group within the development set, to account for class imbalances. All other parameters were set to the default values in the Matlab Classification Learner App.

- Decision trees:
  - Fine (max: 100 splits); medium (max: 20 splits); coarse (max: 4 splits).
- Discriminant analysis:
  - Linear; quadratic.
- Logistic regression.
- Naïve Bayes
  - Gaussian; kernel.
- Support vector machines:
  - Linear; quadratic; cubic; fine Gaussian (Gaussian kernel scale: $\sqrt{p}\ /4$, with P the number of predictors); medium Gaussian (kernel scale: $\sqrt{p}$) coarse Gaussian (kernel scale: $4 * \sqrt{p}$).
- K Nearest Neighbours:
  - Fine KNN (1 neighbour); medium KNN (10 neighbours); coarse KNN (100 neighbours); cosine KNN; cubic KNN; weighted KNN.
- Ensemble Classifiers:
  - Boosted trees (AdaBoost algorithm); bagged trees; subspace discriminant; subspace KNN; RUSBoosted trees.
- Neural networks:
  - Narrow (one fully connected layer of size 10); medium (one fully connected layer of size 25); wide (one fully connected layer of size 100); bilayered; trilayered.

**Model Performance & Outcome Metrics**
**Normal clinically non-informative**

**Predicted Class**

| Clinical Diagnosis | Epilepsy | Differential | |
|---|---|---|---|
| Epilepsy | True Positive: 41 (21%) | False Negative: 26 (13%) | Sensitivity: 61.19% |
| Differential | False Positive: 33 (17%) | True Negative: 97 (49%) | Specificity: 74.62% |
| | Positive Predictive Value: 55.41% | Negative Predictive Value: 78.86% | **Balanced Accuracy:** 67.90% |

- False Negative Rate: 0.3881
- False Positive Rate: 0.2538
- False Discovery Rate: 0.4459
- False Omission Rate: 0.2114
- Positive Likelihood Ratio: 2.4107
- Negative Likelihood Ratio: 0.5201
- Threat Score: 0.4100
- F1 Score: 0.5816
- Matthews Correlation Coefficient: 0.3503
- ROCAUC: 0.7158
- Brier Score: 0.2001

**Abnormal clinically non-informative**

**Predicted Class**

| Clinical Diagnosis | Epilepsy | Differential | |
|---|---|---|---|
| **Epilepsy** | True Positive: 43 (46%) | False Negative: 27 (29%) | Sensitivity: 61.43% |
| **Differential** | False Positive: 9 (10%) | True Negative: 14 (15%) | Specificity: 60.87% |
| | Positive Predictive Value: 82.69% | Negative Predictive Value: 34.15% | **Balanced Accuracy:** 61.15% |

- False Negative Rate: 0.3857
- False Positive Rate: 0.3913
- False Discovery Rate: 0.1731
- False Omission Rate: 0.6585
- Positive Likelihood Ratio: 1.5698
- Negative Likelihood Ratio: 0.6337
- Threat Score: 0.5443
- F1-Score: 0.7049
- Matthews Correlation Coefficient: 0.1938
- ROCAUC: 0.6534
- Brier Score: 0.2554

**Individual Biomarker Trends for the Clinically Non-Informative & Normal Cohort**

- Peak alpha:
  - Original finding: lower in epilepsy.
  - Observed trend: discordant.
- Alpha frequency:
  - Original finding: lower in epilepsy.
  - Observed trend: discordant.
- Mean degree:
  - Original finding: higher in epilepsy.
  - Observed trend: not discordant.
- Degree variance:
  - Original finding: higher in epilepsy.
  - Observed trend: not discordant.
- Clustering coefficient:
  - Original finding: higher in epilepsy.
  - Observed trend: not discordant.
- Characteristic path-length:
  - Original finding: higher in controls.
  - Observed trend: not discordant.
- Critical coupling:
  - Original finding: lower in epilepsy;
  - Observed trend: concordant.
- Local coupling:
  - Original finding: higher order parameter in epilepsy;
  - Observed trend: not discordant.

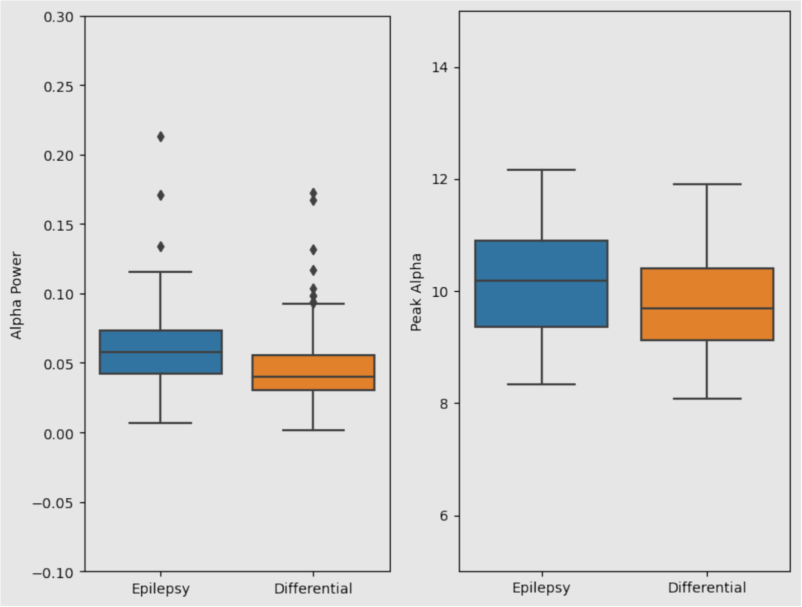

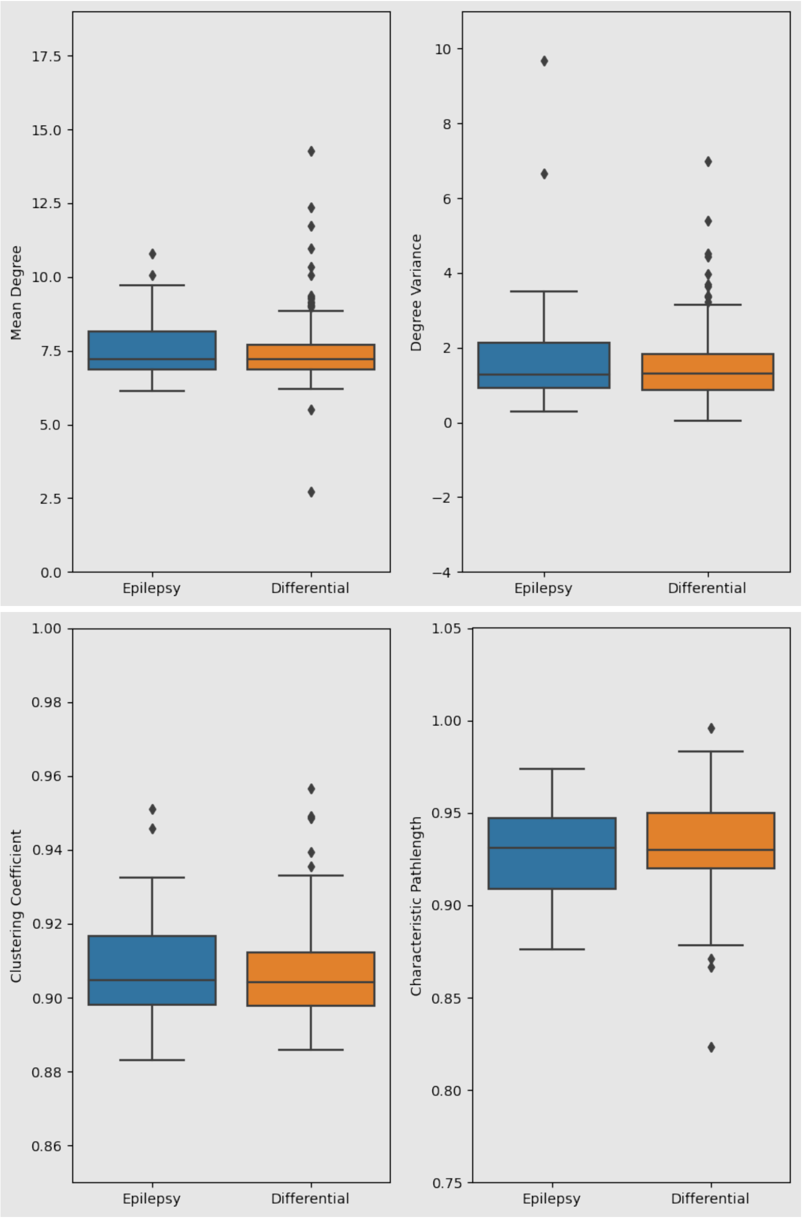
Mean Degree
Degree Variance
Clustering Coefficient
Characteristic Pathlength
Epilepsy
Differential

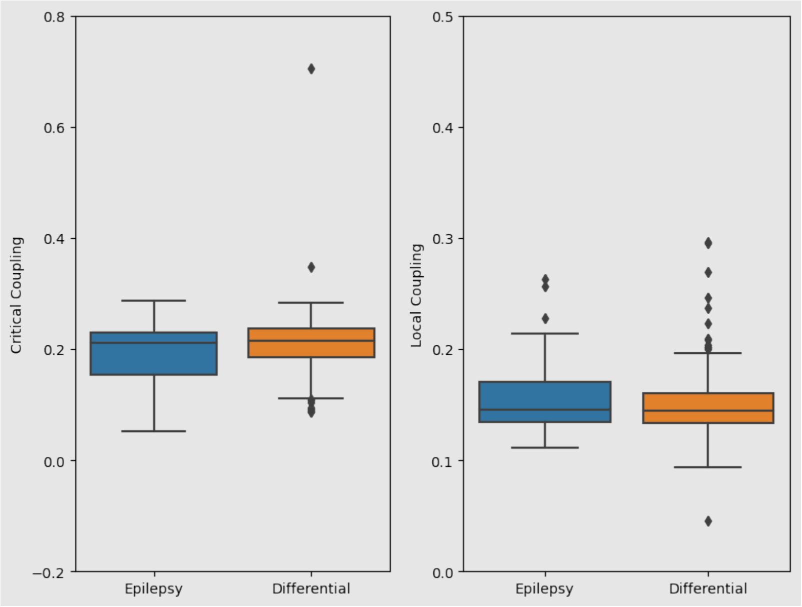
Critical Coupling
0.8
0.6
0.4
0.2
0.0
−0.2
Epilepsy
Differential
Local Coupling
0.5
0.4
0.3
0.2
0.1
0.0
Epilepsy
Differential

**Mistake Analysis**

We used Fisher's exact test (at 95%) to study the potential impact of categorical variables on overall performance, and the Kruskal-Wallis test (95%) for the continuous variables:

**Uncorrected p-values**

Sex: 0.5328
Comorbidity: 0.2311
ASM status: 0.4090
Sites: [0.3323, 0.9314, 0.2524, 0.5104]
Age: 0.5682
